# Sex differences in cardiometabolic traits at four life stages: cohort study with repeated metabolomics

**DOI:** 10.1101/2020.01.15.19015206

**Authors:** Joshua A. Bell, Diana L. Santos Ferreira, Abigail Fraser, Ana Luiza G. Soares, Laura D. Howe, Deborah A. Lawlor, David Carslake, George Davey Smith, Linda M. O’Keeffe

**Affiliations:** MRC Integrative Epidemiology Unit at the University of Bristol, Bristol, UK; Population Health Sciences, Bristol Medical School, University of Bristol, Bristol, UK; Bristol NIHR Biomedical Research Centre, Bristol, UK; School of Public Health, Western Gateway Building, University College Cork, Ireland

**Keywords:** Sex differences, Cardiometabolic traits, Coronary heart disease, NMR metabolomics, Epidemiology, ALSPAC

## Abstract

**Background:** Males experience higher rates of coronary heart disease (CHD) than females, but the circulating traits underpinning this difference are poorly understood. We examined sex differences in detailed cardiometabolic traits measured at four life stages, spanning childhood to middle adulthood.

**Methods and Results:** Data were from the Avon Longitudinal Study of Parents and Children cohort study. 229 traits quantified from targeted metabolomics (nuclear magnetic resonance spectroscopy) including lipoprotein subclass-specific cholesterol and triglycerides, amino acids, glucose, and inflammatory glycoprotein acetyls were measured repeatedly in offspring (Generation 1 (G1)) born in 1991-92 and once in their parents (Generation 0 (G0)). Measurements in G1 were once in childhood (mean age 8y), twice in adolescence (16y and 18y) and once in early adulthood (25y), and in G0 once in middle adulthood (50y). Linear regression models were used to examine differences in standardized traits for males compared with females on each occasion (serial cross-sectional associations). 7,727 G1s (49% male) and 6,500 G0s (29% male) contributed to analyses. At age 8y, total lipids in very-low-density lipoproteins (VLDL) were lower in males than females; levels were higher in males than females at age 16y and were higher still by age 18y and age 50y (in G0) for medium-or-larger subclasses. Larger sex differences at older ages were most pronounced for triglycerides in VLDL – e.g. male levels were 0.19 standard deviation (SD) units (95% CI=0.12, 0.26) higher at age 18y, 0.50 SD (95% CI=0.42, 0.57) higher at age 25y, and 0.62 SD (95% CI=0.55, 0.68) higher at age 50y. Cholesterol in VLDL and low-density lipoproteins (LDL) was generally lower in males, with inconsistent sex differences across ages. Apolipoprotein-B was generally lower in males than females. Branched chain amino acids were consistently higher in males after age 8y with the largest sex difference of all traits at all ages seen for leucine at age 50y (1.53 SD, 95% CI=1.47, 1.58 higher in males compared with females). Males had consistently lower glycoprotein acetyls across ages.

**Conclusions:** Our results suggest that males begin to have higher VLDL triglycerides in adolescence, and that this sex difference is larger at older ages. Sex differences in other CHD-related traits, including LDL cholesterol, apolipoprotein-B, and inflammatory glycoproteins, show the opposite pattern with age, with higher levels among females. Higher triglyceride content may therefore be a key factor underpinning the higher age-adjusted rate of CHD among males; causal analyses of this and other traits are needed to understand whether they differentially affect CHD risk among males and females.

## Introduction

Coronary heart disease (CHD) remains the leading cause of death globally (1, 2). Recent decades have seen age-adjusted incidence and mortality rates decline substantially in higher income countries (3), but those declines are now slowing (4, 5) and total numbers of cases are rising in most countries owing to population ageing and growth (2, 6). Age-adjusted CHD rates are higher among males than females (7), and reasons for this are gaining clarity. For example, males are known to store more fat in visceral and ectopic compartments which drives insulin resistance (8, 9) and results in higher type 2 diabetes rates among males (10). Males also have higher systolic blood pressure than females from adolescence to middle adulthood, although this difference narrows or even reverses in older age (11-13).

Sex differences in circulating lipid traits are more contradictory. Adult females tend to have lower triglyceride levels compared with adult males, potentially due to hormonal mechanisms (14, 15), yet adult females also tend to have higher cholesterol in low-density lipoprotein (LDL) particles (12, 16). Such comparisons have been based mostly on circulating traits measured by conventional clinical assays. More detailed measures from targeted metabolomic platforms are now available (17) which have helped characterise the cardiometabolic profile of pregnancy and menopause among females (18, 19). Knowledge of sex differences in these more detailed traits at multiple life stages may help reveal more specific circulating pathways that underpin sex differences in age-adjusted rates of CHD, but no such investigation has yet been conducted.

We aimed in this study to better characterise sex differences in CHD-relevant cardiometabolic traits at multiple life stages. Using data from a multi-generational pregnancy cohort study, we examined mean differences by sex in over 200 circulating cardiometabolic traits quantified from targeted metabolomics, including lipoprotein subclass-specific cholesterol and triglycerides, amino acids and inflammatory traits, at four life stages. These traits were measured once in childhood (mean age 8y), twice in adolescence (16y and 18y), and once in early adulthood (25y) on both male and female participants (Generation 1 (G1)), as well as on their parents (Generation 0 (G0)) in middle adulthood (age 50y).

## Methods

### Study population

Data were from participants of the Avon Longitudinal Study of Parents and Children (ALSPAC), a population-based birth cohort study in which 14,541 pregnant women expected to deliver between 1 April 1991 and 31 December 1992 were recruited from the former county of Avon in southwest England. Offspring (G1 cohort) alive at 1 year (n=13,988) have since been followed up with multiple assessments with an additional 913 children enrolled over the course of the study (20, 21). Mothers and fathers (termed hereafter as ‘partners’ as not all are biological fathers) of offspring participants have also been followed with multiple assessments (G0 cohort) (22). Parental data used here were primarily from mothers who attended a clinic assessment between December 2008 and July 2011 and from partners who attended a clinic assessment between September 2011 and February 2013.

Ethical approval was obtained from the ALSPAC Law and Ethics and Local Research Ethics Committees. The study website contains details of all the data that is available through a fully searchable data dictionary and variable search tool (http://www.bristol.ac.uk/alspac/researchers/our-data/).

### Assessment of cardiometabolic traits

Among offspring (G1 cohort), blood samples were drawn in clinics at mean (standard deviation, SD) ages 7.5y (0.3y), 15.4y (0.3y), 17.7y (0.4y), and 24.5y (0.8y). Proton nuclear magnetic resonance (^1^H-NMR) spectroscopy from a targeted metabolomics platform (23) was performed on EDTA-plasma samples from each of these four occasions to quantify 229 traits (149 concentrations plus 80 ratios) including cholesterol, triglyceride, and other lipid content in lipoprotein subclass particles, apolipoproteins, fatty acids and amino acids, and inflammatory glycoprotein acetyls. Bloods at age 8y were taken while not fasting and bloods at age 16y, 18y, and 25y were taken after a minimum of a 6-hour fast (stability in these trait concentrations has been shown over different fasting durations (24)).

Among parents (G0 cohort), blood samples were drawn in clinics at mean (SD) age 47.9y (4.5y) among mothers and 53.3y (5.4y) among partners. The same ^1^H-NMR metabolomics platform used among G1 offspring was performed on serum samples taken on G0 parents after a minimum of a 6-hour fast during these clinics to quantify the same 229 traits described above. Among mothers only, additional blood samples were available from visits conducted between July 2011 to June 2013 when mothers were of mean (SD) age 50.8y (4.4y), from which additional measures of the same 229 cardiometabolic traits were quantified with NMR as prior. If mothers were missing data on all traits on the first measurement occasion but had data on at least one trait on the second measurement occasion, then trait values from that second occasion were used to replace missing values on the first occasion. The number of mothers with these replacements ranged from 215 to 223 across traits.

### Participants eligible for analyses

To allow full use of measured data, analyses were conducted using maximum numbers of participants (with N varying across ages and between traits). Participants were eligible for inclusion in analyses at any age if they had data on sex, age, and at least one of the cardiometabolic traits. This resulted in 7,727 eligible G1 offspring (3,763 males, 3,964 females) contributing to some analyses, with sample sizes for age-specific analyses ranging from 5,403 to 5,515 at age 8y, from 3,162 to 3,358 at age 16y, from 3,090 to 3,174 at age 18y, and from 3,204 to 3,260 at age 25y. The single time-point analysis in G0 parents included 6,500 eligible parents (1,855 males, 4,645 females), with sample sizes for each trait ranging between 5,800 and 6,500. No specific exclusions were made based on cardiometabolic diagnoses or medication use in G1 or G0.

### Statistical approach

We examined characteristics of participants eligible for inclusion in analyses by sex; these included age at each occasion, ethnicity, highest level of maternal and partner education at the time of G1 offspring birth, prevalence of maternal and partner smoking during pregnancy and their pre-pregnancy body mass index (BMI), BMI of the G1 offspring and G0 parents as measured at clinics, puberty timing of G1 offspring (age at peak height velocity), and menopausal status of G0 mothers at the time cardiometabolic traits were measured (determined via questionnaire using the Stages of Reproductive Age Workshop (STRAW) criteria (18, 25)); the measurement of all these characteristics are described further in **Supplementary Methods**. These characteristics were also examined among a group of participants who were not eligible for inclusion in our analyses, defined as not having data on sex, age, and at least 1 cardiometabolic trait at any measurement occasion (6,085 G1 offspring and 7,811 G0 parents) to examine the potential for selection bias. We additionally examined the above-mentioned characteristics in G0 females, G1 males and G1 females by the participation status of G0 males (mothers’ partners), to assess potential selection bias.

Cardiometabolic traits at each measurement occasion were standardized into SD units using z-scores (subtracting the sex-combined mean and dividing by the sex-combined SD). Linear regression models with robust standard errors (to accommodate skewed outcome distributions) were used to examine the mean difference and 95% confidence interval (CI) for the association of sex with each standardized cardiometabolic trait on each occasion, adjusting for age at the time of trait assessment. In light of differences in the age distribution of G0 parents, we centred age at 50y and included an interaction term between sex and centred age to allow associations of sex with cardiometabolic traits to differ by age. Estimates are therefore interpreted as the difference in mean (in SD units) of each cardiometabolic trait for males compared with females. All models were additionally run using original (non-SD; mostly mmol/l) units to aid clinical interpretation. Meta-regression was used to examine whether effect estimates change linearly with mean age on each occasion, via P-values for trend across occasions. This was applied for purposes of directly testing heterogeneity in estimates across offspring and parents (analysed separately) under an independence assumption.

### Supplementary analyses

The main analyses of sex differences in G0 parents (when mean age was 47.9y among mothers and 53.3y among partners) include females who are predominantly pre-menopause (62.1% of females reported current menstrual periods). To better examine the impact of natural menopause on results, we re-examined sex differences in cardiometabolic traits among G0 parents when including, first: only those females who were considered to be pre-menopause; and second: only those females who were considered to be post-menopause (i.e. ≥ 1 year post-menopause and neither pre-menopause nor peri-menopause as defined as menopause transition; described in **Supplementary Methods**). Both analyses excluded females who reported having either a hysterectomy, oophorectomy, ablation/resection, chemotherapy or radiation therapy (i.e. surgical menopause), use of hormonal contraception, or use of hormone replacement (18, 25).

All models were repeated on a complete case sample of 769 G1 offspring (46% male) and 5,187 G0 parents (19% male) who had complete data on sex, age, and every cardiometabolic trait on every occasion. This was done to ensure that results were not driven by the changing sample across time points.

Since our statistical aims involve estimation, we present exact P-values without imposing arbitrary significance thresholds and focus on effect size and precision (26-28). Analyses were conducted using Stata 15.1 (StataCorp, College Station, Texas, USA).

## Results

### Sample characteristics

Male and female G1 offspring had a similar mean age on each occasion – mean (SD) age overall was 7.5y (0.3y), 15.5y (0.3y), 17.8y (0.4y), and 24.5y (0.8y) successively (**Table 1**). A minority of males and females (each < 5.0%) were of a non-white ethnicity. Maternal and partner education levels were similar among male and female G1 offspring, as was the prevalence of maternal and partner smoking during/around the time of pregnancy. Mean (SD) age at peak height velocity among G1 offspring was 12.6y (1.3y) overall; this was later among males than females at 13.6y (0.9y) and 11.7y (0.8y), respectively. Based on this indicator, 4.5% of male and 5.7% of female G1 offspring were post-pubertal on the first measurement occasion (at 8y; i.e. had reached their peak height prior to the 8y clinic), while 100% of females and 97% of males were post-pubertal on the second measurement occasion (at 16y; i.e. had reached their peak height prior to the 16y clinic).

**Table 1.** Characteristics of ALSPAC G1 offspring and G0 parents eligible for analyses

| **Characteristics** | **G1 Offspring** |  |  |  | **G0 Parents** |  |  |  |
| --- | --- | --- | --- | --- | --- | --- | --- | --- |
|  | **N** | **Males** | **N** | **Females** | **N** | **Males** | **N** | **Females** |
| Age (y) on first clinic occasion – mean (SD) | 3410 | 7.5 (0.3) | 3376 | 7.5 (0.3) | 1855 | 53.2 (5.3) | 4645 | 47.9 (4.5) |
| Non-white ethnicity – % (N) | 3328 | <5.0 (NA*) | 3422 | <5.0 (NA*) | 4175 | <5.0 (NA*) | 4219 | <5.0 (NA*) |
| Highest maternal education – % (N) | 3389 |  | 3470 |  | - |  | 4233 |  |
| Certificate of Secondary Education |  | 13.3 (450) |  | 14.0 (484) |  | - |  | 10.2 (430) |
| Vocational |  | 8.4 (286) |  | 8.4 (293) |  | - |  | 7.4 (313) |
| Ordinary level |  | 35.7 (1209) |  | 34.2 (1185) |  | - |  | 34.3 (1453) |
| Advanced level |  | 26.3 (891) |  | 26.9 (932) |  | - |  | 29.3 (1238) |
| Degree |  | 16.3 (553) |  | 16.6 (576) |  | - |  | 18.9 (799) |
| Highest partner education – % (N) | 3293 |  | 3378 |  | 4142 |  | - |  |
| Certificate of Secondary Education |  | 19.0 (625) |  | 21.5 (725) |  | 16.9 (699) |  | - |
| Vocational |  | 7.6 (251) |  | 7.7 (259) |  | 7.4 (308) |  | - |
| Ordinary level |  | 22.3 (734) |  | 21.4 (723) |  | 21.0 (869) |  | - |
| Advanced level |  | 28.1 (925) |  | 27.4 (926) |  | 29.3 (1213) |  | - |
| Degree |  | 23.0 (758) |  | 22.1 (745) |  | 25.4 (1053) |  | - |
| BMI on first clinic occasion – mean (SD) | 3385 | 16.1 (1.9) | 3350 | 16.4 (2.2) | 1831 | 27.5 (4.0) | 4629 | 26.6 (5.3) |
| Maternal BMI pre-pregnancy – mean (SD) | 3163 | 22.9 (3.8) | 3246 | 22.8 (3.6) | - | - | 3991 | 22.5 (3.4) |
| Partner BMI at pregnancy – mean (SD) | 2438 | 25.1 (3.1) | 2544 | 25.2 (3.3) | 1493 | 24.7 (3.0) | - | - |
| Maternal smoking during pregnancy – % (N) | 3133 | 22.5 (706) | 3181 | 22.4 (713) | - | - | 3919 | 17.0 (665) |
| Partner smoking during pregnancy – % (N) | 3271 | 29.7 (972) | 3333 | 31.6 (1054) | 4070 | 27.3 (1112) | - | - |
| Age (y) at peak height velocity – mean (SD) | 2404 | 13.6 (0.9) | 2688 | 11.7 (0.8) | - | - | - | - |
| Menopause status (STRAW) – % (N) | - | - | - | - | - | - | 3,513 |  |
| Pre-menopause |  | - |  | - |  | - |  | 62.1 (2,181) |
| Peri-menopause |  | - |  | - |  | - |  | 19.4 (680) |
| Post-menopause |  | - |  | - |  | - |  | 18.6 (652) |

Male G0 parents (partners) were on average older than female G0 parents (mothers) at the time of assessment – mean (SD) age was 53.2y (5.3y) among males, vs 47.9y (4.5y) among females (**Table 1**). A similarly low proportion of males and females (each < 5.0%) were of a non-white ethnicity, while more males than females reported a degree as their highest educational qualification (25.4% vs. 18.9% respectively). Smoking at any time during the pregnancy was more common among males than females (27.3% vs. 17.0%, respectively). Among mothers, 62.1% were considered to be pre-menopause by the time of clinic assessment; whereas 19.4% were peri-menopause and 18.6% were post-menopause.

Participants described are those with data on sex, age, and at least 1 cardiometabolic trait on any measurement occasion. ‘Maternal/partner’ characteristics refer to own status among parents. BMI: Body mass index. STRAW: Stages of Reproductive Age Workshop. *Cells have been censored due to small cell counts in accordance with study ethics.

As shown in **Supplementary Table 1**, G1 offspring who were ineligible for any analysis had a lower parental educational attainment than those who were eligible, and were more likely to have parents who reported smoking during pregnancy – e.g. maternal smoking during pregnancy was 42.3% among male offspring who were ineligible, vs. 22.5% among those who were eligible. These patterns were also seen among G0 parents themselves, with lower education and higher levels of smoking during pregnancy among participants ineligible for analyses – e.g. 34.6% of ineligible mothers smoked, compared with 17.0% of eligible mothers. As shown in **Supplementary Table 2**, G0 mothers whose partner did participate were similar to mothers whose partner did not participate in terms of maternal age, ethnicity, BMI (current and pre-pregnancy), and menopause status; but mothers whose partner did participate had a higher proportion of degree holders (25.2% vs 15.5%) and a lower proportion of smoking during pregnancy (11.1% vs 20.5%) compared with mothers whose partners did not participate. As shown in **Supplementary Table 3**, G1 participants whose mothers’ partners did participate were similar to G1 participants whose mothers’ partners did not participate in terms of age, BMI (measured and maternal/paternal pre-pregnancy), and puberty timing; but G1 participants whose mothers’ partners did participate were less likely to be of a non-white ethnicity, more likely to have parents with academic degrees, and less likely to have parents who smoked during pregnancy. The magnitude of the difference by sex in these characteristics among G1 offspring did not differ greatly by mothers’ partners participation status.

### Sex differences in lipid traits

At age 8y, total lipids were lower among males than females in all lipoprotein subclasses including very-low-density lipoproteins (VLDL), intermediate-density lipoproteins (IDL), and low-density lipoproteins (LDL), except for high-density lipoprotein (HDL) subclasses in which total lipids were higher among males (full results in **Supplementary Table 4**). At age 16y, levels of total lipids in (medium and larger) VLDL subclasses were similar between the sexes, but sex differences in these emerged at age 18y (among G1 offspring) and were evident at age 50y (among G0 parents) for subclasses that were medium or larger – e.g. total lipids in large VLDL were higher among males than females by 0.21 SD (95% CI = 0.14, 0.28), by 0.45 SD (95% CI = 0.37, 0.52), and by 0.72 SD (95% CI = 0.65, 0.79) at ages 18y, 25y, and 50y, respectively. P-values for trend across occasions based on meta-regressions were generally lowest for lipids in VLDL, supporting patterns of higher levels at older ages – e.g. P=0.02 for total lipids in large VLDL. The higher levels of VLDL lipids seen among males at older ages were most pronounced for triglyceride content in VLDL (**Figure 1**). Cholesterol was higher among males than females in large VLDL particles, but lower among males in other particles including LDL, with inconsistent sex differences at age 25y apart from cholesterol in HDL (**Figure 2**). Sex differences in lipoprotein particle sizes themselves were larger at older ages – appearing higher among males for VLDL and lower among males for HDL (**Figure 3**). Apolipoprotein-B was also notably lower among males than females at all ages apart from 25y, while apolipoprotein-B as a function of apolipoprotein-A-1 was higher among males at older ages (**Figure 3**).

**Figure 1.**
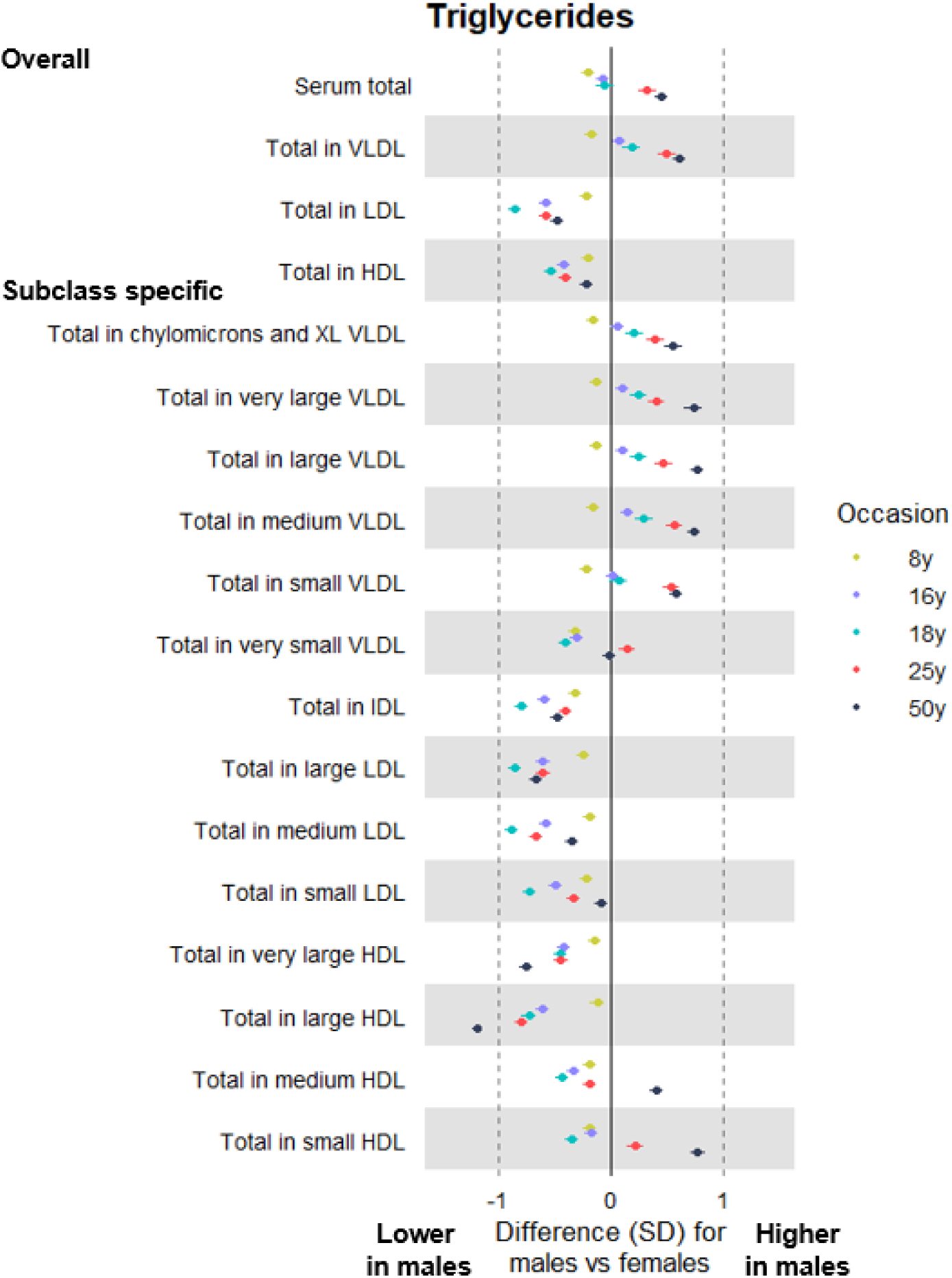
Sex differences in levels of lipoprotein triglyceride content at different life stages among ALSPAC G1 offspring and G0 parents **Legend** Trait measures at mean age 8y, 16y, 18y and 25y are among ALSPAC G1 offspring; trait measures at mean age 50y are among ALSPAC G0 parents (analysed separately). VLDL: Very-low-density lipoprotein. IDL: Intermediate-density lipoprotein. LDL: Low-density lipoprotein. HDL: High-density lipoprotein.

**Figure 2.**
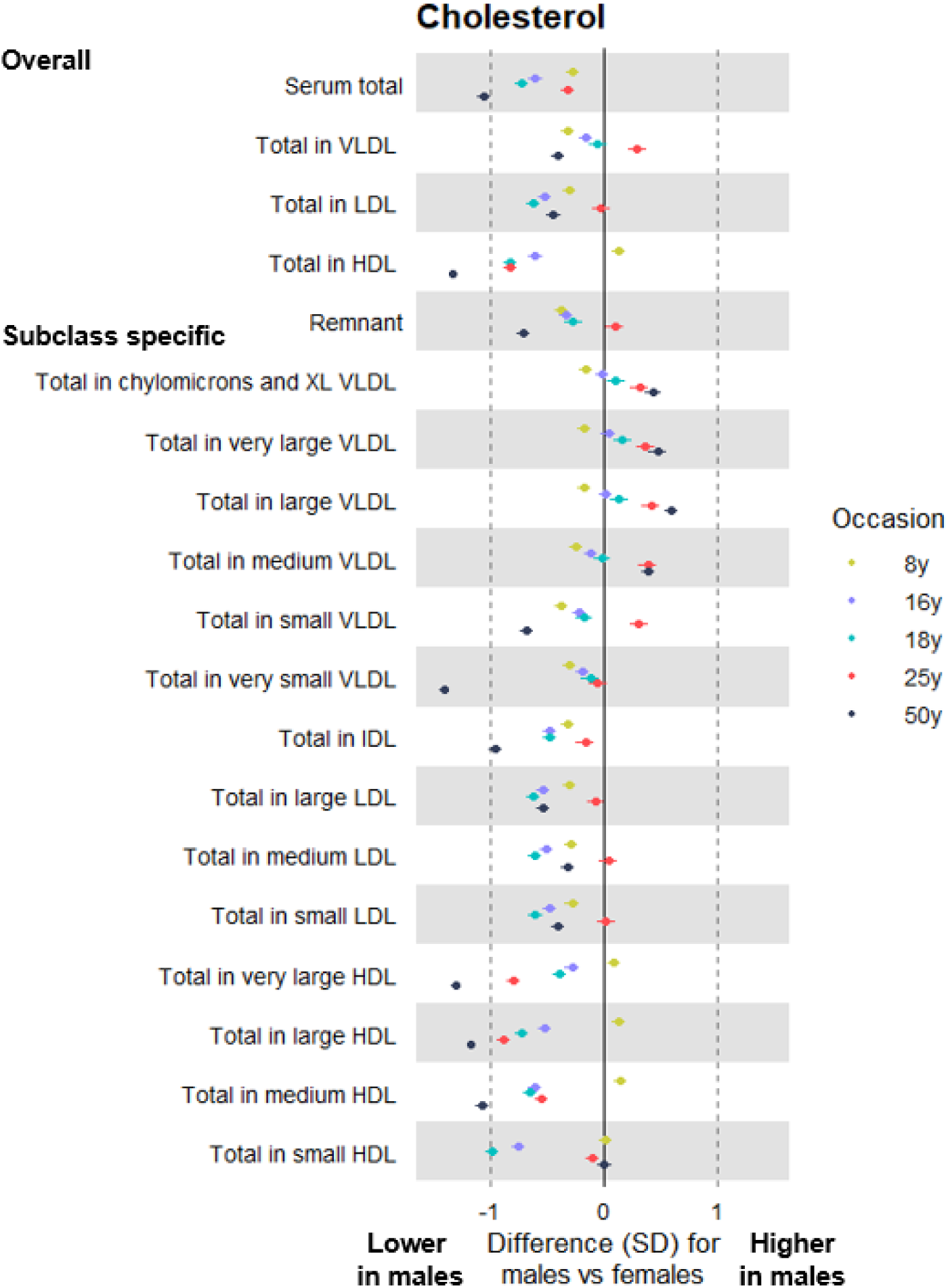
Sex differences in levels of lipoprotein cholesterol content at different life stages among ALSPAC G1 offspring and G0 parents **Legend** Trait measures at mean age 8y, 16y, 18y and 25y are among ALSPAC G1 offspring; trait measures at mean age 50y are among ALSPAC G0 parents (analysed separately). VLDL: Very-low-density lipoprotein. IDL: Intermediate-density lipoprotein. LDL: Low-density lipoprotein. HDL: High-density lipoprotein.

**Figure 3.**
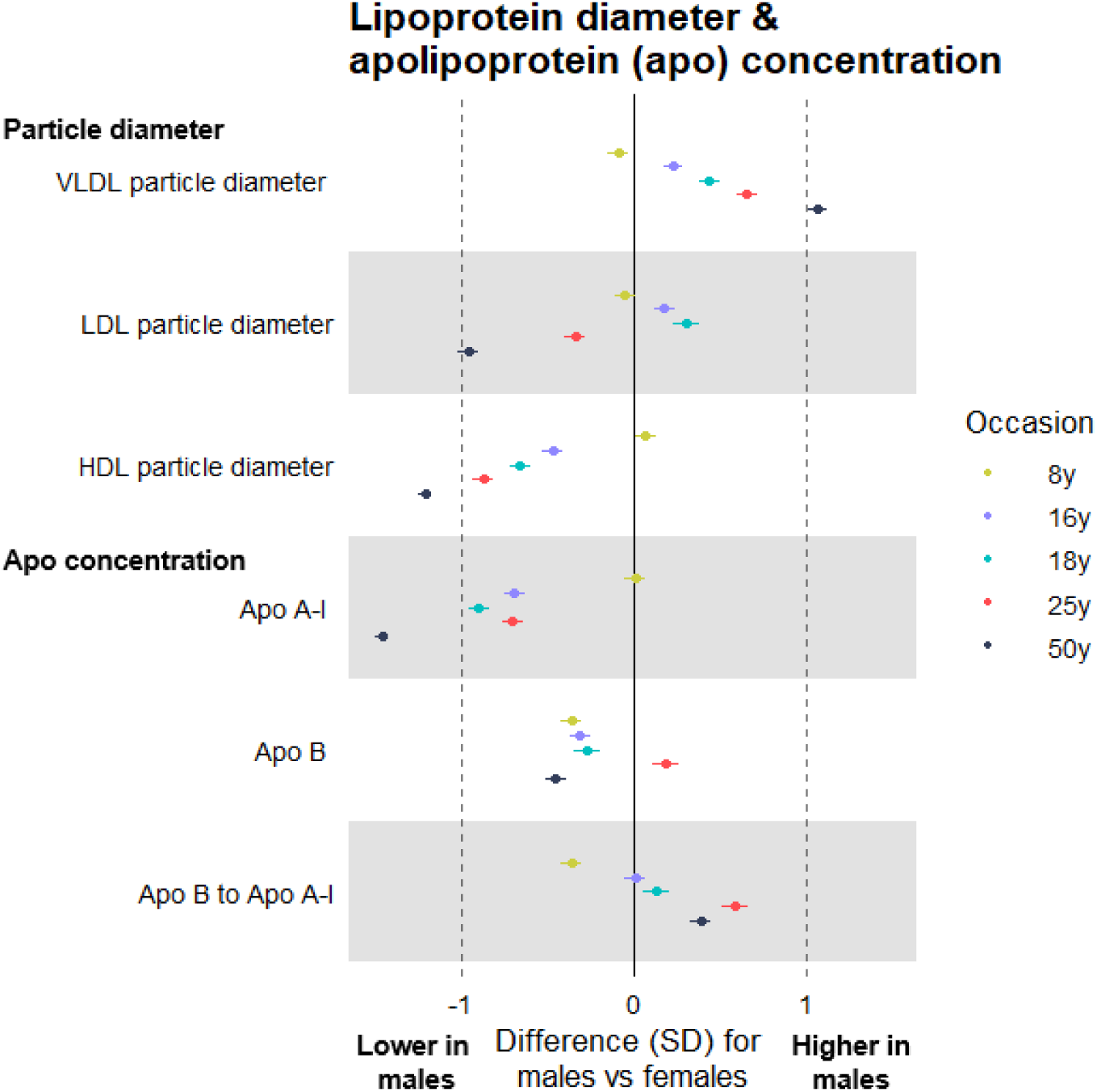
Sex differences in levels of lipoprotein particle size and apolipoprotein concentration at different life stages among ALSPAC G1 offspring and G0 parents **Legend** Trait measures at mean age 8y, 16y, 18y and 25y are among ALSPAC G1 offspring; trait measures at mean age 50y are among ALSPAC G0 parents (analysed separately). VLDL: Very-low-density lipoprotein. LDL: Low-density lipoprotein. HDL: High-density lipoprotein.

### Sex differences in glycemic and inflammatory traits

Fatty acid levels tended to be lower among males than females, with some evidence that this difference was larger at older ages – e.g. males had lower polyunsaturated fatty acids by −0.26 SD (95% CI = −0.32, −0.21) at age 8y among G1 offspring and by −1.10 SD (95% CI = −1.16, −1.05) at age 50y among G0 parents (**Figure 4**; full results in **Supplementary Table 4**). Glucose was consistently higher among male G1 offspring, whereas among G0 parents at age 50y glucose was −0.24 SD (95% CI = −0.30, −0.17) lower among males. Lactate and citrate levels were markedly higher among males at age 50y, with sex differences of 0.73 SD (95% CI = 0.66, 0.79) and 1.28 SD (95% CI = 1.23, 1.32), respectively. Amino acids were consistently higher among males after age 8y, particularly branched chain amino acids – e.g. leucine was 0.06 SD (95% CI = 0.003, 0.11) higher among males at age 8y and 1.53 SD (95% CI = 1.47, 1.58) higher among males at age 50y. Comparably large sex differences were seen at age 50y for isoleucine (1.29 SD, 95% CI = 1.23, 1.34 higher among males) and for the aromatic amino acid phenylalanine (1.44 SD, 95% CI = 1.40, 1.49 higher among males).

**Figure 4.**
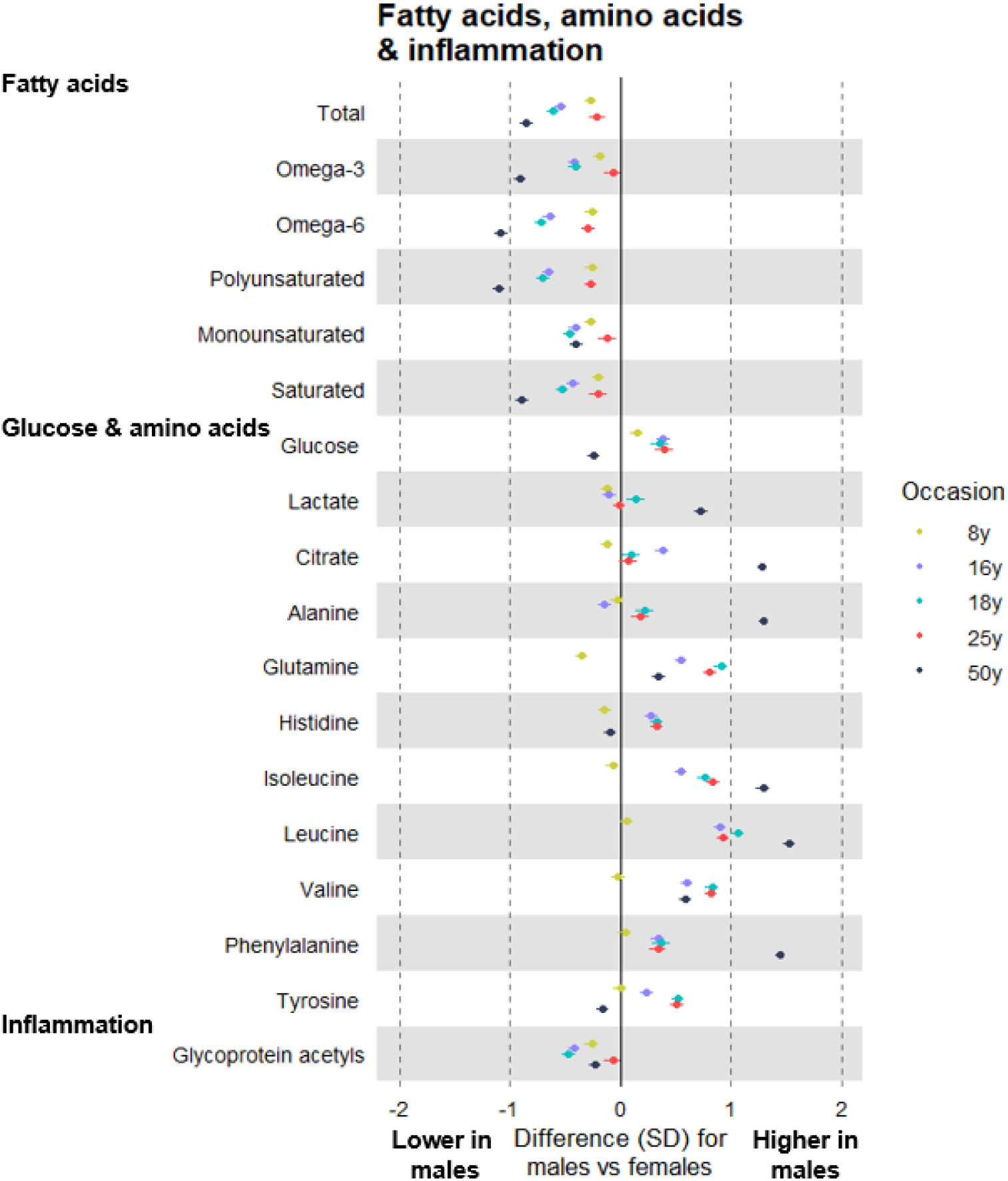
Sex differences in levels of fatty acids, glycolysis-related metabolites, amino acids, and glycoprotein acetyls at different life stages among ALSPAC G1 offspring and G0 parents **Legend** Trait measures at mean age 8y, 16y, 18y and 25y are among ALSPAC G1 offspring; trait measures at mean age 50y are among ALSPAC G0 parents (analysed separately).

Small sex differences were seen in levels of ketone bodies at older ages. Creatinine levels were consistently higher among males than females after age 8y; this difference was largest at age 25y (1.24 SD, 95% CI = 1.18, 1.30) and was smaller at age 50y (0.51 SD, 95% CI = 0.45, 0.56; **Supplementary Table 4**). Males had consistently lower inflammatory glycoprotein acetyl levels with little evidence of trend across occasions (meta-regression P-value = 0.62); the magnitude of this difference was smaller after age 18y (**Figure 4**).

Full results of above sets of analyses using original (non-SD) units are shown in **Supplementary Table 5**.

### Supplementary analyses

When comparing middle-aged males with females who were pre-menopause, sex differences in traits were similar to main estimates (full results in **Supplementary Table 6;** full results in non-SD units in **Supplementary Table 7**). When comparing middle-aged males with females who were post-menopause, sex differences in traits were of a lower magnitude, particularly for lipids in VLDL – e.g. total lipids in large VLDL were 0.66 SD (95% CI = 0.58, 0.73) higher among males. In contrast, sex differences were larger for lipids in IDL, LDL, and HDL particles – e.g. total cholesterol in IDL was −1.01 SD (95% CI = −1.08, - 0.95) lower among males. Sex differences were lower for branched chain amino acids – e.g. valine was 0.53 SD (95% CI = 0.47, 0.60) higher among males. In contrast, sex differences were generally larger for fatty acid traits, glucose, and glycoprotein acetyls – e.g. −0.36 SD (95% CI = −0.44, −0.29) lower among males for glycoprotein acetyls.

Sex differences were similar when above sets of analyses were repeated using a complete case sample of 769 G1 offspring and 5,187 G0 parents with data on every cardiometabolic trait on every measurement occasion (full results in **Supplementary Table 4-7**), with expectedly lower precision given reduced statistical power.

## Discussion

In this study, we examined sex differences in detailed cardiometabolic traits at multiple life stages, from childhood to middle adulthood, to help identify circulating traits that may underpin sex differences in CHD burden. Our results suggest that, from adolescence onwards, triglycerides in VLDL are higher among males while levels of other CHD-related traits including LDL cholesterol, apolipoprotein-B, and inflammatory glycoprotein acetyls, are higher among females. Higher triglyceride content may therefore be a key factor underpinning the higher age-adjusted rate of CHD among males; causal analyses of this and other traits are needed to understand whether they differentially affect CHD risk among males and females.

A causal role for LDL cholesterol in the aetiology of CHD is strongly supported by human genetic (29, 30) and pharmacological intervention (31, 32) studies. Recent genetic evidence also supports a role of higher apolipoprotein-B in CHD independent of LDL cholesterol concentration (33-36). Despite this, our results suggest that differences in absolute levels of LDL cholesterol or apolipoprotein-B are unlikely to underpin the higher risk of CHD experienced among males, since levels were lower (more favourable) among males in adolescence and young adulthood in the G1 offspring cohort, as well as in middle adulthood in the G0 parent cohort. However, this does not exclude the possibility of LDL cholesterol, apolipoprotein-B, or other traits which did not differ by sex in these analyses having a differential effect on CHD among males and females; sex-stratified causal analyses of these in relation to CHD itself are needed to determine this. The present study addressed absolute effects of sex on cardiometabolic traits, not relative effects of cardiometabolic traits on CHD by sex.

In contrast, our results suggest that triglyceride content – particularly in VLDL particles – are higher (more adverse) among males in adolescence in the offspring cohort and that this sex difference is larger in young adulthood, and larger still among males in middle adulthood in the parent cohort. The tendency for males to have higher circulating triglycerides has been observed previously (14, 15), but how differences relative to females progress across multiple life stages has been poorly understood due to lack of repeated measures. Higher triglyceride concentrations among males at multiple life stages observed here (in both the G1 offspring and G0 parent cohort), together with previous genetic evidence of a causal role of triglycerides for CHD (34, 37), support triglycerides as a key target for CHD prevention, particularly among males. Whether triglycerides increase CHD risk more greatly among males requires sex-stratified analyses.

Equally strong tendencies were found for lower HDL cholesterol among males than females at later life stages. However, despite robust observational associations of lower HDL cholesterol with higher CHD risk (38), genetic (29, 39) and pharmacological intervention studies (40) do not support a causal effect of HDL cholesterol on CHD. Nevertheless, HDL cholesterol is increasingly regarded as a useful non-causal marker for insulin resistance and other traits that are causal, namely circulating triglycerides, with which it tracks strongly in opposing directions (38, 41). The utility of HDL cholesterol as a marker of pre-glycemic changes is further supported by results here showing progressively lower levels of HDL cholesterol to coincide with progressively higher levels of branched chain amino acids, which are likely components of early-stage insulin resistance (42).

The earlier measurement occasions used in this study (age 8y and 16y) spanned puberty for most participants – an important transitional period of growth and development. The mean age at puberty onset here was estimated at 13.6y for males and 11.7y for females based on growth-curve modelling of repeated height measures. This transition profoundly influences physical and sexual maturity (43, 44), and among females results in the release of estrogen and other sex hormones which are thought to result in a less adverse lipid profile (15); whether testosterone release among males itself results in a more adverse lipid profile is less clear (15, 45). Potential beneficial effects of estrogen or adverse effects of testosterone on lipid profiles are supported by present results which suggest that total lipids and triglycerides in VLDL are lower among males before puberty, but then switch direction after the onset of puberty to become higher among males in adolescence and young adulthood, a difference which is even greater in middle adulthood. This was also true of cholesterol in HDL particles but in the reverse direction (higher levels among males before puberty, then lower levels after puberty). Together, this supports puberty as a pivotal time for the emergence of life-long sex differences in triglyceride and risk-marking HDL cholesterol levels.

How these sex differences extend beyond middle age, when the menopausal transition among females is complete, is less clear. Females in middle adulthood were measured here at mean age 47.9y, when natural menopause has either not yet begun or is typically in its early stages. Menopause status was examined here using the rigorous STRAW criteria (18, 25), and sex differences in cardiometabolic traits were re-examined when including only those females who were considered to be pre-menopause; these indicated similar results as seen when all females were included. Sex differences were then re-examined when including only those females who were considered to be post-menopause (also excluding those who were peri-menopause), and sex differences in VLDL lipids appeared moderately narrower than were seen when all females were included. In contrast, sex differences in IDL, LDL, and HDL lipids appeared wider. Differences also appeared narrower in branched chain amino acids, but wider in most other traits including fatty acids and glucose. This suggests that the proposed cardio-protective effects of female estrogen release (14, 15), which would be reduced post-menopause (18, 25), are VLDL-specific, but further studies in females with natural and surgical menopause are needed to confirm these patterns.

### Strengths and limitations

Key strengths of this study include its use of five measures of over 200 detailed cardiometabolic traits from a targeted metabolomics platform measured at multiple life stages (four measures in the G1 offspring cohort and one measure in the G0 parent cohort). This offers rare insight into how sex differences in key CHD-related traits track over time, from childhood to middle-age, but also insights into whether the pubertal and menopausal transitions alter these sex differences. The NMR metabolomics platform used here offers extensive coverage of cholesterol and triglyceride content in distinct subclasses of lipoproteins, which enables greater specificity of sex differences in cardiometabolic traits than previously possible.

Limitations of this study include modest sample sizes among G1 offspring participants, particularly for complete case analyses, and greater loss to follow-up among males. Unequal numbers of males and females may result in biased estimates of sex differences if loss to follow-up is related to both sex and outcomes; however, demographic characteristics were comparable between included and excluded males and females, suggesting that such bias is unlikely or small. Estimates were based on serial cross-sectional associations (based on linear regression models at each time point), which did not account for correlation between repeated measures of cardiometabolic traits over time. The data used for analyses were of a unique multi-generational nature, comprising G1 offspring and their G0 parents, but this does open the possibility of additional sources of bias from cohort and period effects. Selection bias could also be differential between G1 offspring and G0 parents. This is indicated by higher proportions of maternal smoking among the included G1 offspring than among the included G0 parents who subsequently participated in clinic assessments several years after pregnancy (e.g. maternal smoking was 22.4% among participating female G1 offspring vs. 17.0% among participating G0 mothers themselves), indicating that mothers who attended clinic assessments years after pregnancy were a relatively healthy subset of those initially recruited. Sex is an essentially randomized exposure within a causal framework with little-to-no expected influence of common causes (confounders), although such factors could influence study participation. Numerous explanations exist for sex differences in cardiometabolic traits described here, and these would largely be considered mediators of associations. Future studies could examine these pathways using formal mediation analyses.

### Conclusions

Measures of detailed cardiometabolic traits at multiple life stages suggest that VLDL triglyceride content is higher among males relative to females in adolescence, and that this difference is larger at older ages. In contrast, other CHD-related traits including LDL cholesterol, apolipoprotein-B, and inflammatory glycoprotein acetyls are higher among females in adolescence, with similar patterns with advancing age. Higher triglyceride content may therefore be a key factor underpinning the higher age-adjusted rate of CHD among males; causal analyses of this and other traits are needed to understand whether they differentially affect CHD risk among males and females.

## Data Availability

Individual-level ALSPAC data are available following an application. This process of managed access is detailed at www.bristol.ac.uk/alspac/researchers/access. Cohort details and data descriptions are publicly available at the same web address.

## Acknowledgments

We are extremely grateful to the families who participated in this study, the midwives for their help in recruiting them, and the whole ALSPAC team which includes interviewers, computer and laboratory technicians, clerical workers, research scientists, volunteers, managers, receptionists, and nurses. The UK Medical Research Council, Wellcome (102215/2/13/2), and the University of Bristol provide core support for ALSPAC. A comprehensive list of grants funding is available on the ALSPAC website (http://www.bristol.ac.uk/alspac/external/documents/grant-acknowledgements.pdf); this research (collection of NMR metabolomics data) was specifically funded by the MRC (MC_UU_12013/1) and NIHR (NF-SI-0611-10196). This publication is the work of the authors who are guarantors for its contents. JAB is supported by Cancer Research UK (C18281/A19169) and the Elizabeth Blackwell Institute for Health Research, University of Bristol and the Wellcome Trust Institutional Strategic Support Fund. AF, ALGS and LDH are funded by UK Medical Research Council research fellowships (MR/M009351/1 & MR/M020894/1, respectively). DAL’s contribution to this work is supported by the European Research Council under the European Union’s Seventh Framework Programme (FP/2007-2013) / ERC Grant Agreement (Grant number 669545; DevelopObese), the European Union’s Horizon 2020 research and innovation programme under grant agreement No 733206 (LifeCycle), the United States National Institutes of Health (NIH): National Institute of Diabetes and Digestive and Kidney Diseases (R01 DK10324), the British Heart Foundation (AA/18/7/34219) and National Institute of Health Research ((NF-SI-0611-10196); LMOK is supported by a Health Research Board (HRB) of Ireland Emerging Investigator Award (EIA-2019-007) and a UK Medical Research Council Population Health Scientist fellowship (MR/M014509/1); DLSF, DAL, DC and GDS work in a unit funded by the UK Medical Research Council (MC_UU_00011/1,6) and the University of Bristol. The funders had no role in study design, data collection and analysis, decision to publish, or preparation of the manuscript.

## References

1. Naghavi M, Abajobir AA, Abbafati C, Abbas KM, Abd-Allah F, Abera SF, et al. Global, regional, and national age-sex specific mortality for 264 causes of death, 1980–2016: a systematic analysis for the Global Burden of Disease Study 2016. Lancet. 2017;390(10100):1151–210.

2. Roth GA, Johnson C, Abajobir A, Abd-Allah F, Abera SF, Abyu G, et al. Global, regional, and national burden of cardiovascular diseases for 10 causes, 1990 to 2015. J Am Coll Cardiol. 2017;70(1):1–25.

3. Unal B, Critchley JA, Capewell S. Explaining the decline in coronary heart disease mortality in England and Wales between 1981 and 2000. Circulation. 2004;109(9):1101–7.

4. Wilmot KA, O’flaherty M, Capewell S, Ford ES, Vaccarino V. Coronary heart disease mortality declines in the United States from 1979 through 2011: evidence for stagnation in young adults, especially women. Circulation. 2015;132(11):997–1002.

5. Sidney S, Quesenberry CP, Jaffe MG, Sorel M, Nguyen-Huynh MN, Kushi LH, et al. Recent trends in cardiovascular mortality in the United States and public health goals. JAMA Cardiol. 2016;1(5):594–9.

6. Roth GA, Forouzanfar MH, Moran AE, Barber R, Nguyen G, Feigin VL, et al. Demographic and epidemiologic drivers of global cardiovascular mortality. N Engl J Med. 2015;372(14):1333–41.

7. Mosca L, Barrett-Connor E, Kass Wenger N. Sex/gender differences in cardiovascular disease prevention: what a difference a decade makes. Circulation. 2011;124(19):2145–54.

8. Kahn SE, Hull RL, Utzschneider KM. Mechanisms linking obesity to insulin resistance and type 2 diabetes. Nature. 2006;444(7121):840–6.

9. Rosen ED, Spiegelman BM. What we talk about when we talk about fat. Cell. 2014;156(1):20–44.

10. Kautzky-Willer A, Harreiter J, Pacini G. Sex and gender differences in risk, pathophysiology and complications of type 2 diabetes mellitus. Endocrine Rev. 2016;37(3):278–316.

11. Wills AK, Lawlor DA, Matthews FE, Sayer AA, Bakra E, Ben-Shlomo Y, et al. Life course trajectories of systolic blood pressure using longitudinal data from eight UK cohorts. PLoS Med. 2011;8(6):e1000440.

12. O’Keeffe LM, Simpkin AJ, Tilling K, Anderson EL, Hughes AD, Lawlor DA, et al. Sex-specific trajectories of measures of cardiovascular health during childhood and adolescence: A prospective cohort study. Atherosclerosis. 2018;278:190–6.

13. O’Keeffe LM, Simpkin AJ, Tilling K, Anderson EL, Hughes AD, Lawlor DA, et al. Data on trajectories of measures of cardiovascular health in the Avon Longitudinal Study of Parents and Children (ALSPAC). Data in Brief. 2019;23:103687.

14. Link JC, Reue K. Genetic basis for sex differences in obesity and lipid metabolism. Ann Rev Nutr. 2017;37:225–45.

15. Palmisano BT, Zhu L, Eckel RH, Stafford JM. Sex differences in lipid and lipoprotein metabolism. Mol Metab. 2018;15:45.

16. Moran A, Jacobs Jr DR, Steinberger J, Steffen LM, Pankow JS, Hong C-P, et al. Changes in Insulin Resistance and Cardiovascular Risk During Adolescence. Establishment of Differential Risk in Males and Females. Circulation. 2008;117(18):2361–8.

17. Würtz P, Kangas AJ, Soininen P, Lawlor DA, Davey Smith G, Ala-Korpela M. Quantitative Serum NMR Metabolomics in Large-Scale Epidemiology: A Primer on-Omic Technology. Am J Epidemiol. 2017:kwx016.

18. Wang Q, Ferreira DLS, Nelson SM, Sattar N, Ala-Korpela M, Lawlor DA. Metabolic characterization of menopause: cross-sectional and longitudinal evidence. BMC Med. 2018;16(1):17.

19. Wang Q, Würtz P, Auro K, Mäkinen V-P, Kangas AJ, Soininen P, et al. Metabolic profiling of pregnancy: cross-sectional and longitudinal evidence. BMC Med. 2016;14(1):205.

20. Boyd A, Golding J, Macleod J, Lawlor DA, Fraser A, Henderson J, et al. Cohort profile: the ‘children of the 90s’—the index offspring of the Avon Longitudinal Study of Parents and Children. Int J Epidemiol. 2012(1):111–27.

21. Northstone K, Lewcock M, Groom A, Boyd A, Macleod J, Timpson N, et al. The Avon Longitudinal Study of Parents and Children (ALSPAC): an update on the enrolled sample of index children in 2019. Wellcome Open Res. 2019;4.

22. Fraser A, Macdonald-Wallis C, Tilling K, Boyd A, Golding J, Davey Smith G, et al. Cohort profile: the Avon Longitudinal Study of Parents and Children: ALSPAC mothers cohort. Int J Epidemiol. 2013;42(1):97–110.

23. Soininen P, Kangas AJ, Würtz P, Suna T, Ala-Korpela M. Quantitative serum nuclear magnetic resonance metabolomics in cardiovascular epidemiology and genetics. Circ Cardiovasc Genet. 2015;8(1):192–206.

24. Sidhu D, Naugler C. Fasting time and lipid levels in a community-based population: a cross-sectional study. Arch Intern Med. 2012;172(22):1707–10.

25. Harlow SD, Gass M, Hall JE, Lobo R, Maki P, Rebar RW, et al. Executive summary of the Stages of Reproductive Aging Workshop+ 10: addressing the unfinished agenda of staging reproductive aging. J Clin Endocrinol Metab. 2012;97(4):1159–68.

26. Sterne JA, Davey Smith G. Sifting the evidence—what’s wrong with significance tests? BMJ. 2001;322(7280):226–31.

27. Wasserstein RL, Lazar NA. The ASA’s statement on p-values: context, process, and purpose. Am Statistician. 2016;70(2):129–33.

28. Amrhein V, Greenland S, McShane B. Scientists rise up against statistical significance. Nature; 2019.

29. Holmes MV, Asselbergs FW, Palmer TM, Drenos F, Lanktree MB, Nelson CP, et al. Mendelian randomization of blood lipids for coronary heart disease. Eur Heart J. 2014;36(9):539–50.

30. Ference BA, Majeed F, Penumetcha R, Flack JM, Brook RD. Effect of naturally random allocation to lower low-density lipoprotein cholesterol on the risk of coronary heart disease mediated by polymorphisms in NPC1L1, HMGCR, or both: a 2× 2 factorial Mendelian randomization study. J Am Coll Cardiol. 2015;65(15):1552–61.

31. Sabatine MS, Giugliano RP, Keech AC, Honarpour N, Wiviott SD, Murphy SA, et al. Evolocumab and clinical outcomes in patients with cardiovascular disease. N Engl J Med. 2017;376(18):1713–22.

32. Cannon CP, Blazing MA, Giugliano RP, McCagg A, White JA, Theroux P, et al. Ezetimibe added to statin therapy after acute coronary syndromes. N Eng J Med. 2015;372(25):2387–97.

33. Ference BA, Kastelein JJ, Ginsberg HN, Chapman MJ, Nicholls SJ, Ray KK, et al. Association of genetic variants related to CETP inhibitors and statins with lipoprotein levels and cardiovascular risk. JAMA. 2017;318(10):947–56.

34. Ference BA, Kastelein JJ, Ray KK, Ginsberg HN, Chapman MJ, Packard CJ, et al. Association of triglyceride-lowering LPL variants and LDL-C–lowering LDLR variants with risk of coronary heart disease. JAMA. 2019;321(4):364–73.

35. Sniderman AD. Type III Hyperlipoproteinemia: The Forgotten, Disregarded, Neglected, Overlooked, Ignored but Highly Atherogenic, and Highly Treatable Dyslipoproteinemia. Clin Chem; 2019.

36. Sniderman A, Couture P, De Graaf J. Diagnosis and treatment of apolipoprotein B dyslipoproteinemias. Nat Rev Endocrinol. 2010;6(6):335.

37. Do R, Willer CJ, Schmidt EM, Sengupta S, Gao C, Peloso GM, et al. Common variants associated with plasma triglycerides and risk for coronary artery disease. Nat Genet. 2013;45(11):1345.

38. Collaboration PS. Blood cholesterol and vascular mortality by age, sex, and blood pressure: a meta-analysis of individual data from 61 prospective studies with 55 000 vascular deaths. Lancet. 2007;370(9602):1829–39.

39. Voight BF, Peloso GM, Orho-Melander M, Frikke-Schmidt R, Barbalic M, Jensen MK, et al. Plasma HDL cholesterol and risk of myocardial infarction: a mendelian randomisation study. Lancet. 2012;380(9841):572–80.

40. Schwartz GG, Olsson AG, Abt M, Ballantyne CM, Barter PJ, Brumm J, et al. Effects of dalcetrapib in patients with a recent acute coronary syndrome. N Eng J Med. 2012;367(22):2089–99.

41. Khera AV, Kathiresan S. Genetics of coronary artery disease: discovery, biology and clinical translation. Nat Rev Genet. 2017;18(6):331.

42. Wang Q, Holmes MV, Davey Smith G, Ala-Korpela M. Genetic support for a causal role of insulin resistance on circulating branched-chain amino acids and inflammation. Diab Care. 2017:dc171642.

43. Day FR, Bulik-Sullivan B, Hinds DA, Finucane HK, Murabito JM, Tung JY, et al. Shared genetic aetiology of puberty timing between sexes and with health-related outcomes. Nat Commun. 2015;6:8842.

44. Perry JR, Murray A, Day FR, Ong KK. Molecular insights into the aetiology of female reproductive ageing. Nat Rev Endocrinol. 2015(11):725–34.

45. Fernández-Balsells MM, Murad MH, Lane M, Lampropulos JF, Albuquerque F, Mullan RJ, et al. Adverse effects of testosterone therapy in adult men: a systematic review and meta-analysis. J Clin Endocrinol Metab. 2010;95(6):2560–75.

